# Emotional Processing After Severe Traumatic Brain Injury: Insights from Functional MRI and Pupillometry

**DOI:** 10.1101/2025.08.13.25333244

**Authors:** Karnig Kazazian, Stefanie O. Zhang, Jack de Jeu, Phoebe K. Lawrence, William R. Sanders, Anogue Meydan, Fion Bremner, Emma-Jane Mallas, Yelena G. Bodien, Kristen Dams-O’Connor, Brian L. Edlow, Lucia M. Li

## Abstract

**Objective:** Emotional dysfunction is a common consequence of severe traumatic brain injury (TBI), yet the mechanisms underlying these symptoms remain poorly understood. This study investigated whether brain network and autonomic mechanisms involved in emotional processing are abnormal in TBI.

**Methods:** We conducted a cross-sectional study of chronic severe TBI (n=26) and healthy control participants (n=15). We analysed functional MRI (fMRI) data to assess brain processing of emotionally salient music (joyful and fearful stimuli; n=15 TBI, n=15 controls), and resting-state fMRI (rsfMRI) to measure the functional connectivity of relevant intrinsic brain networks (limbic, salience, and default mode networks; n=16 TBI, n=15 controls). We additionally measured the pupillary light reflect (PLR) to assess parasympathetic and sympathetic function (n=14 TBI, n=11 controls).

**Results:** Individuals with severe TBI did not demonstrate the left insula activation elicited by joyful versus fearful musical stimuli seen in healthy controls. rsfMRI revealed decreased connectivity between the salience network, caudate and hippocampus in severe TBI compared to controls. Exploratory analyses identified reduced connectivity between default mode (bilateral medial prefrontal cortex) and limbic (bilateral amygdala) nodes in TBI compared to controls. PLR measurements revealed blunted dark-adaptation responses in individuals with severe TBI compared to controls (F(1,24)=27.4, p<0.001).

**Interpretation:** Individuals with chronic severe TBI show reduced insula activation during emotional stimuli processing, resting connectivity abnormalities in salience, limbic and default mode networks, and evidence of sympathetic dysfunction. Brain network and autonomic alterations may be potential neural mechanisms of post-TBI emotional dysregulation.

## Introduction

Traumatic brain injury (TBI) is a significant global health problem, with approximately 70 million people affected worldwide every year.^1^ Neurobehavioral disturbances can have the most devastating impact on quality of life through disruption of an individual’s sense of self, relationships, and social functioning.^2^ Rates of mood and anxiety disorders, emotional lability, apathy, irritability and aggression, and suicide are markedly increased after TBI.^3,4^ Despite the significant impact of post-TBI neurobehavioral sequelae, there is a gap in our understanding of the pathophysiologic mechanisms that underly these clinical manifestations, limiting the development of targeted interventions to improve symptoms.^2^

Neuroimaging studies have demonstrated brain network dysfunction and abnormalities in dopaminergic transmission in TBI survivors with depression.^5–7^ Notably, these changes appear to be distinct from those observed in depression arising from other causes, suggesting that we cannot simply extrapolate our understanding of TBI-related neurobehavioral mechanisms from the general psychiatry literature.^6,8^ Moreover, prior brain network studies in patients with post-traumatic psychological symptoms have focused on resting-state analyses, leaving it unclear how the observed network disruptions contribute to emotional function. A transdiagnostic approach to that integrates stimulus-based and resting-state analyses may offer greater mechanistic insight by identifying core component deficits (e.g., aberrant emotional stimuli processing and emotional recognition) that contribute to heterogeneous clinical presentations.^9^ This approach has proven useful in understanding cognitive dysfunction after TBI.^10^

Behavioral studies in individuals with TBI suggest that abnormalities in emotional recognition and processing may contribute to impairments in social function post-TBI.^11–14^ Music can evoke strong emotions, and prior functional MRI (fMRI) studies show that processing of emotional music stimuli in healthy adults reliably causes activation in limbic regions, including the amygdala and insula.^15^ Further, music stimuli elicit distinct patterns of neural activation in these regions depending on their emotional valence, reflecting the role of these regions in emotional valence processing. Assessing how an individual’s brain responds to emotionally evocative stimuli may thus help identify neural mechanisms of emotional processing deficits in individuals with TBI.

In this study, we recruited individuals with chronic severe TBI enrolled in the Late Effects of Traumatic Brain Injury (LETBI) study.^16^ We used stimulus-based fMRI, comprised of musical stimuli to elicit feelings of joy and fear, to investigate emotional processing post-TBI. Resting-state fMRI was used to investigate functional connectivity differences between individuals with TBI and healthy controls in intrinsic brain networks that are relevant for emotional processing: the salience (SN) and default mode networks (DMN). Disruptions in structural and functional connectivity within both SN and DMN have been associated with a broad range of neuropsychological and cognitive impairments after TBI.^17,18^

Further, autonomic dysfunction may be another important contributing mechanism to neurobehavioral symptoms after TBI.^19^ Prior work has demonstrated evidence of both parasympathetic and sympathetic dysfunction in the chronic phase of TBI.^19^ Given the strong bidirectional links between autonomic function and emotional processing, particularly through shared circuitry involving the insula and amygdala,^20–22^ we sought to complement our neuroimaging data with an objective measure of autonomic function. This was acquired through quantitative assessment of the pupillary response to luminance changes.

Using this trans-diagnostic approach, we hypothesized that (1) activation in the posterior insula and amygdalae, regions previously shown to differentiate musical emotional stimuli, are altered in individuals with chronic severe TBI compared to controls; (2) within- and between-network connectivity of the SN and DMN are disrupted compared to controls; and (3) there is evidence of both parasympathetic and sympathetic autonomic dysfunction in chronic severe TBI, which relates to the brain activation and connectivity changes.

## Methods

This study is reported in accordance with the Strengthening the Reporting of Observational Studies in Epidemiology (STROBE) guidelines for reporting observational studies.^23^

### Participant Demographics

#### TBI patients

26 individuals with chronic severe TBI were enrolled in the LETBI study^16,24^ at Massachusetts General Hospital in Boston, United States between 2021and 2022. Participants in the current study were a subset of larger LETBI cohort who met the following inclusion criteria: i) history of severe TBI, as defined by a total Glasgow Coma Scale (GCS) score of 8 or less at the time of ICU admission; ii) at least 12 months from TBI; and iii) at least 18 years of age. Exclusion criteria were i) history of neurological or psychiatric disorder prior to the TBI; and ii) contraindication to MRI.

#### Healthy Controls

15 age- and sex-matched healthy adult individuals were recruited as controls. Exclusion criteria were i) history of psychological or neurological disorders, ii) history of cardiovascular, pulmonary, renal or endocrine disorder; iii) current use of psychotropic drugs and iv), alcohol abuse.

### Experimental Protocol

Participants underwent MRI and assessments of level of consciousness and autonomic function. Prior to fMRI acquisition, the consciousness level of each patient was assessed using the Coma Recovery Scale-Revised (CRS-R) and Confusion Assessment Protocol (CAP).^25,26^ Autonomic assessment was performed using pupillometry. Anxiety and depression symptoms were assessed using Quality of Life in Neurological Disorders (Neuro-QOL) short forms in patients who were able to provide self-report.^27^ Data collection were performed following the National Institutes of Health Common Data Element Guidelines for TBI.^28^

### Ethics Statement

Participants or their legal representatives provided written informed consent in accordance with the experimental protocol approved by the Mass General Brigham Institutional Review Board.

### Stimulus-based fMRI Block Design

MRI acquisition parameters are outlined in Supporting Information. Extracts of music were used to test emotional processing of fear and joy.^29^ Stimuli were obtained from a study investigating emotional processing (available online at https://www.stefan-koelsch.de/stimulus_repository/joy_fear_neutral_music.zip). The stimuli were previously rated by healthy participants for their emotional valence, arousal, joy, and fear levels, confirming that each stimulus was perceived as intended.^29^ Stimuli did not have lyrics or fade-in-fade-out sections, and they were matched for amplitude. Fear stimuli included soundtracks of horror series, while joy stimuli featured various musical styles, including jazz and classical music. Neutral stimuli were non-melodic arrangements of musical notes. The experimental paradigm followed a block design (Fig 1), with all stimuli presented once across two separate scanning runs. See supporting information for further methodological details.

**FIGURE 1.**
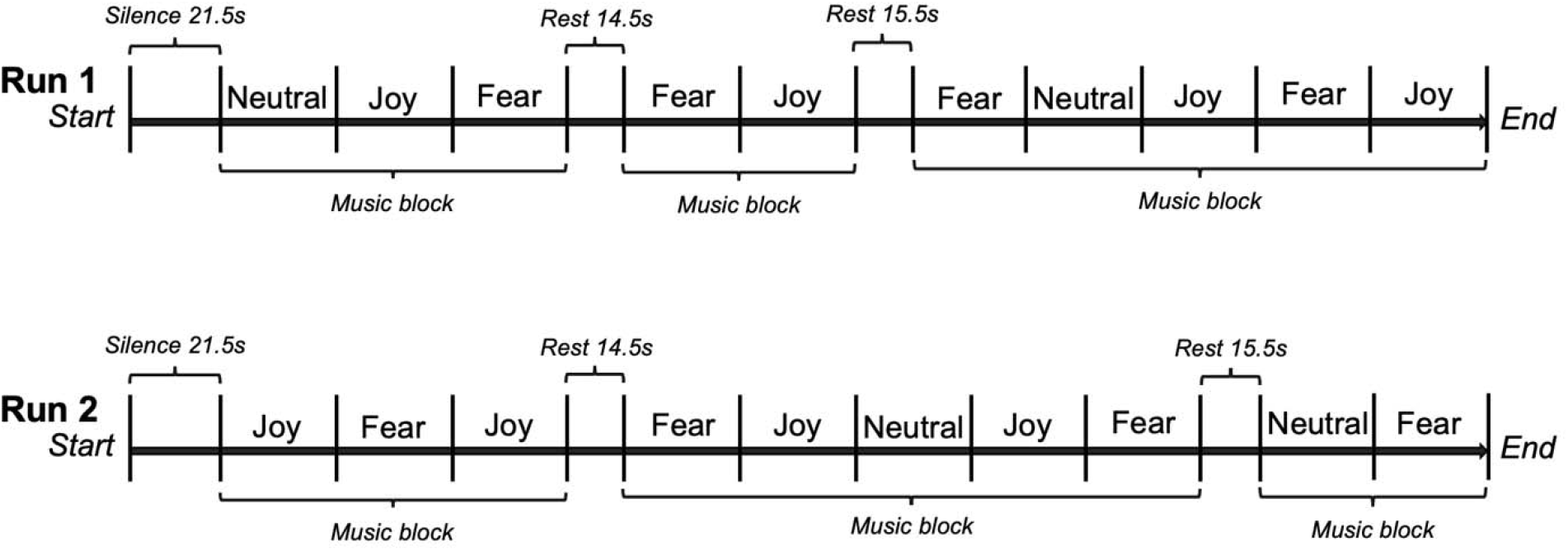
Experiment Paradigm. Each participant underwent two scanning runs. For each scanning run, 10 music stimuli (either joy, fear, or neutral) were pseudorandomly presented across three music blocks, each consisting of different numbers of 30.25s music stimuli, separated by two rest periods of 14.5 and 15.5 seconds, respectively.

### Data Analysis: Stimulus-based fMRI

#### Preprocessing

fMRI data was pre-processed using FMRI Expert Analysis Tool (FEAT) Version 6.00, which is part of the FMRIB’s Software Library (FSL, www.fmrib.ox.ac.uk/fsl.^30,31^ In brief, the fMRI data were preprocessed using standard pipelines, including motion correction, skull stripping, spatial smoothing, high-pass filtering, and normalization to MNI space. Noise components were removed using ICA-AROMA.

#### Stimulus-based fMRI analysis

Subject-level analysis was performed by constructing general linear models (GLM) with three regressors (fear, joy, neutral) convolved with a boxcar kernel model with canonical double-gamma hemodynamic response function (HRF) using FSL’s FEAT tool. To identify differential patterns of brain activation and functional connectivity associated with distinct emotional valence, rather than just a response to acoustic features of stimuli, emotional contrast of joy>fear was computed. Group-level analysis was performed to combine data from the two scanning runs for each participant, and then the lower-level FEATs across all participants were combined for whole group analyses using FSL’s FMRIB’s Local Analysis of Mixed Effects (FLAME 1+2) analysis.^31,32^ See Supporting Information for further details.

We conducted a whole-brain analysis to identify brain regions that show differential activation during emotional music processing. Following this, we performed a region of interest (ROI) analysis focusing on brain areas previously identified as central to emotional music processing (Fig 2A). Anatomical masks were generated based on prior literature to isolate these regions, specifically targeting the insulae and amygdalae. The insula was selected based on findings from Koelsch et al. (2021), which demonstrated that activation clusters within the right and left posterior insula carried significant information about the difference between joyful and fearful musical stimuli in an fMRI study using passive music stimuli of varying emotional valence.^33^ The posterior insula is also considered a key region for interoception, an important process in emotional processing.^34^ The amygdala was included based on Koelsch et al. (2020), a meta-analysis of fMRI studies identifying brain regions involved in processing emotional music stimuli.^15^=

**FIGURE 2.**
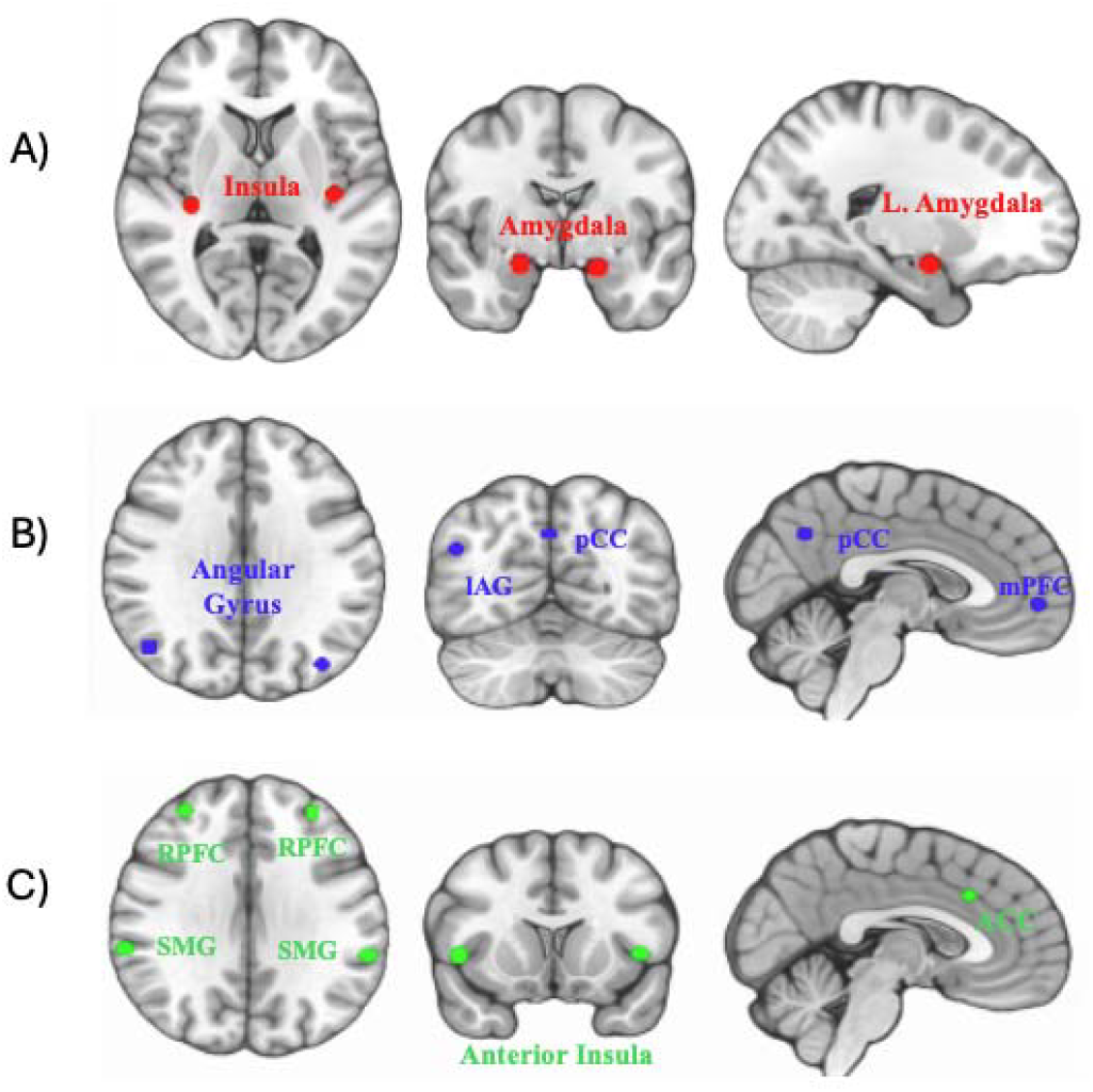
Anatomical Locations of ROIs Used in stimulus and resting-state fMRI Analyses. Axial (left), coronal (middle), and sagittal (right) brain slices showing the anatomical locations of regions of interest (ROIs) included in the stimulus-based and resting-state fMRI analyses. (A) The insula and ACC (limbic network; red) are shown bilaterally on axial and coronal slices, respectively and used for stimulus-based analyses. ROIs were defined as 5 mm spheres centered on coordinates derived from prior literature: right insula (34, -24, 10), left insula (−38, -18, 5), right amygdala (20, -4, -10), left amygdala (−18, -6, -12). (B) The default mode network characterized by: medial prefrontal cortex (mPFC coordinates: 1, 55, -3), left angular gyrus (lAG coordinates: -39, -77, 33), right angular gyrus (rAG coordinates: 47, -67, 29), posterior cingulate cortex (pCC coordinates: 1, -61, 38). Probabilistic seeds are overlayed on a standard MNI152 2mm template. C) Salience networks characterized in part by anterior cingulate cortex (ACC coordinates: 0, 22, 35), left anterior insula (lAI coordinates: -44, 13, 1) right anterior insula (rAI coordinates: 47, 14, 0), left rostral prefrontal cortex (lRPFC coordinates: -32, 46, 27), right rostral prefrontal cortex (rRPFC coordinates: 32, 46, 27), left superior marginal gyrus (lSMG coordinates: -60, -39, 31), and right superior marginal gyrus (rSMG coordinates: 62, -35, 32). SN and DMN used for resting-srtate fMRI analyses.

Separated left and right ROIs were created for structures with bilateral co-ordinates. Spheres (5mm radius) were created using these extracted coordinates and then transformed from Montreal Neurological Institute (MNI) standard space to each participant’s native space for analysis using FSL FMRIB’s Linear Image Registration Tool (FLIRT).^35,36^ Z-statistic images were thresholded using clusters determined by Z>3.1, with no further correction. To identify brain regions underlying differential emotional processing, rather than just a response to acoustic features of stimuli, values were extracted from the [joy>fear] contrast. Group differences between controls and patients in these values were then examined using Welch’s t-test and Mann-Whitney U test, as appropriate.

### Resting State Analysis

Resting-state fMRI data were analyzed using the CONN functional connectivity toolbox (version 22.a) in SPM (version 12.7771).^37^ Preprocessing details are outlined in supporting information.

#### Seed-Based Analysis

Functional connectivity was analyzed using a seed-based approach, where the mean time series in a seed region is compared with the time series of all other voxels in the brain. Anatomical seeds were used to examine the DMN (Figure 2B) and SN (Figure 2C). Detailed information is provided in Supporting Information. Pearson’s correlation coefficients were then calculated between the mean time series and that of each voxel in the brain for each participant, followed by Fisher Z-transformation.

The following group-level differences were investigated using a general linear model accounting for motion as a covariate: 1) healthy controls versus all patients with brain injury (including patients with DoC, defined as vegetative state, minimally conscious state, or post-traumatic confusional state), and 2) healthy controls versus non-DoC patients. To calculate global DMN and SN connectivity values for each participant, the mean Z-score was averaged and extracted from the anatomical ROIs. Global connectivity values were then used to examine differences at the group level, as well as univariate modelling. Global connectivity differences between groups were analyzed using an independent samples t-test, with Welch’s t-test used as appropriate.

#### ROI to ROI Analysis

Functional connectivity between pairs of ROIs was assessed by correlating the BOLD time series of ROI pairs. Pearson correlation coefficients were calculated for each pair and Fisher Z-transformed to normalize the values, resulting in a correlation matrix for each subject. Ten ROIs were selected using the Harvard-Oxford atlas in CONN, including regions within the DMN (medial Prefrontal Cortex [mPFC], left angular gyrus [LAG]), right angular gyrus [RAG], and posterior cingulate cortex [PCC]) and within the SN (rostral prefrontal cortex [RPFC], supramarginal gyrus [SMG], left and right insulae, and left and right amygdalae). Group-level connectivity differences across ROIs were then examined using independent samples t-tests on each ROI pair, with Welch’s t-test used as appropriate. Results were not corrected for multiple comparisons due to the exploratory nature of these analyses.

### Pupillometry

#### Pupillary Reactivity

Pupillometry measurements were performed using both the light and dark stimulus protocols to assess various aspects of the PLR, including constriction velocity, dilation velocity, light amplitude, pupil dilation, and dark adaptation. TBI can disrupt brainstem and hypothalamic pathways, impairing pupil regulation.^19^ We measured dark amplitude to assess sympathetic tone, with reduced dilation in darkness suggesting sympathetic dysfunction. PLR latency was used to probe parasympathetic integrity, as delayed constriction may reflect impaired reflex pathways. Constriction dilation velocities and amplitude were included to quantify the dynamics of both branches of the autonomic nervous system. Detailed descriptions of pupillometry acquisition procedures are provided in the supporting materials. Owing to unreliable pupillometry data acquisition in DOC patients, group differences between healthy controls and non-DOC patients were analyzed using an ANCOVA with age as a covariate.

#### NeuroQOL Analysis

To examine the effects of TBI on quality-of-life, we converted depression and anxiety raw domain scores to T-scores using the look-up tables of the NeuroQOL Version 2.0 User Manual (NINDS, 2015). Statistics for the two domains were based on T-scores, where T = 50 represents the average score of the reference population with a standard deviation of 10 (NINDS, 2015). When a participant missed or skipped a question, a prorated raw score was calculated to approximate their domain score, as per the manual. Non-DOC patients were compared to a reference population by comparing their T-scores on the depression and anxiety domains to the reference population average T = 50, using one-sample *t*-tests (H0: μ =L 50). NeuroQOL was not acquired from healthy controls.

## Results

### Patient information

26 individuals with severe TBI were enrolled (mean age = 34.5 ± 15.3 years; age range = 21-73 years; 15 females). Full demographic and clinical information are outlined in supporting documents (Table S1). Of the 26 patients enrolled, 8 were in a DoC at the time of study procedures (n=3 vegetative state, n=4 minimally conscious state, and n=1 post traumatic confusional state based on CRS-R and CAP assessments).^26,38^ The remaining 18 patients were fully conscious at the time of testing. At initial acute hospital admission, all patients were classified as severe TBI, with a median initial Glasgow Coma Scale score of 4. The mean time of study procedures post-injury was 825.52 ± 815.5 days (range = 194-3770).

Stimulus-based fMRI data were collected from 22 patients. Seven datasets were excluded due to signal dropout and MRI distortion from cranioplasty (n=2), ventriculoperitoneal shunts (n=4), or scan coregistration errors (n=1), leaving 15 patients for stimulus-based fMRI analysis. Resting-state fMRI data was collected from 26 patients, with 10 excluded due to signal dropout (n=5), coregistration errors (n=4), or excessive motion (n=1), leaving 16 patients for resting-state fMRI analysis. Pupillometry was collected from 18 patients, but data from DoC patients was excluded due to unreliable readings, leaving 14 patients in the final analysis. Neuropsychological assessments were acquired in 13 patients who were fully conscious.

15 healthy controls (mean age 38.5 ± 16.0 years; 8 females) were enrolled, matching the TBI cohort in age and sex. Pupillometry was collected from 12 of 15 healthy controls. An overview of all data analyzed in this study is provided in supporting documents (Fig S1).

### Music Stimuli Effectively Elicited Auditory Activity

To confirm that each music stimulus (joy and fear) could effectively elicit neural activity, each condition was contrasted against rest and examined using whole brain analysis. Both stimuli revealed significant BOLD activation signals in patients with TBI and controls in bilateral temporal, insular and DMN regions (mPFC, angular gyrus, and pCC), confirming that auditory processing was broadly intact (Fig 3) in both groups.

**FIGURE 3.**
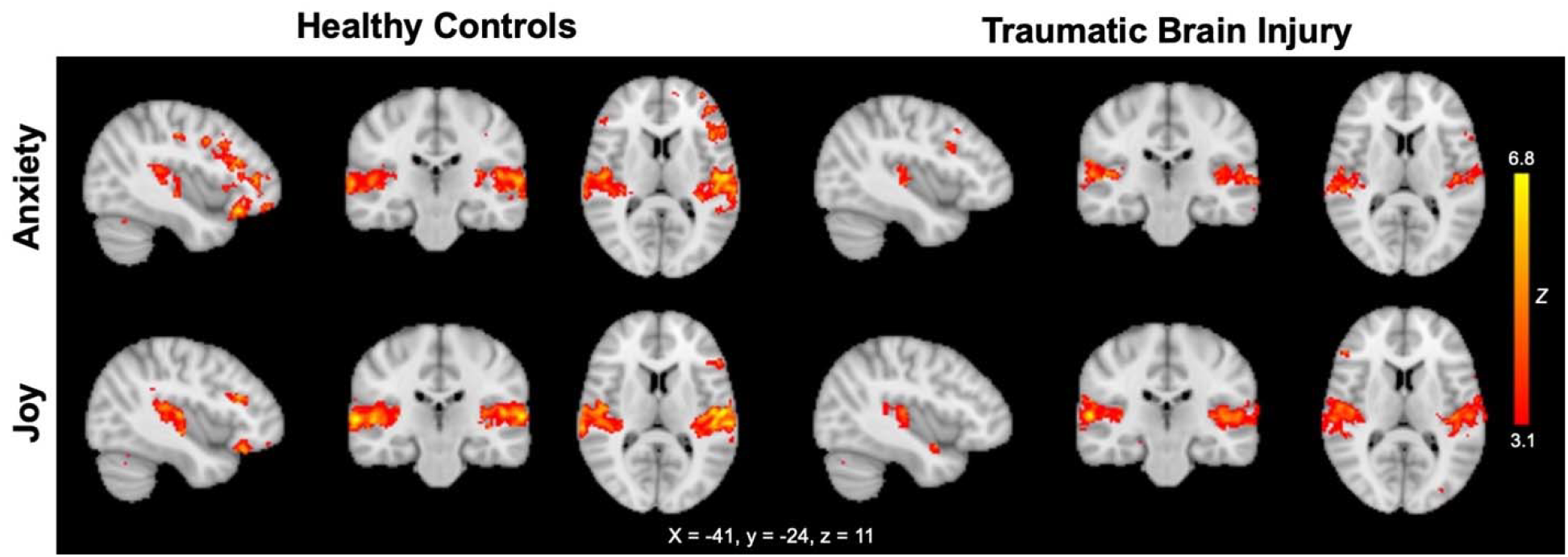
Auditory processing during musical stimuli in controls (n=15) and TBI patients (n=15). Overlay of brain activation associated with each music stimuli of fear and joy, compared to rest, is shown. Z-statistic images were overlayed on the MNI152 2mm standard template and were thresholded using clusters determined by Z>3.1 and a cluster significance threshold of p<0.05.

#### Brain Responses to Joy Compared to Fear

Whole-brain analysis did not reveal any regions showing differential activation between TBI and healthy controls during the [joy>fear] contrast.

In the focused ROI analysis, differences in activation between TBI and healthy controls were identified in the amygdalae and insulae. Specifically, significant group differences were observed in the left insula, where TBI patients (mean activation difference(M)=0.580, SD=5.36) exhibited lower activation than controls (M=6.84, SD=6.69) in response to joy stimuli compared to fear stimuli, T(28)=2.826, p=0.009 (Table 1, Fig 4). This finding was driven by the observation that in TBI patients, left insula activation did not significantly differ between joy and fear stimuli, whereas healthy controls showed greater left insula activation in response to joy compared to fear. This result was significant after Bonferroni correction for multiple comparisons (4 ROIs explored, p_corr_<0.0125), and also after removing DoC TBI patients from the analysis. Non-DoC TBI patients (M=0.28, SD=5.98) demonstrated significantly less activation in the left insula compared to controls (M=6.84, SD=6.69), T(24)=2.58, p=0.016), suggesting that this difference is not simply driven by different arousal levels between the TBI and healthy cohort (Table S3). No group differences in responses within the right insula or amygdalae were identified for this contrast.

**FIGURE 4.**
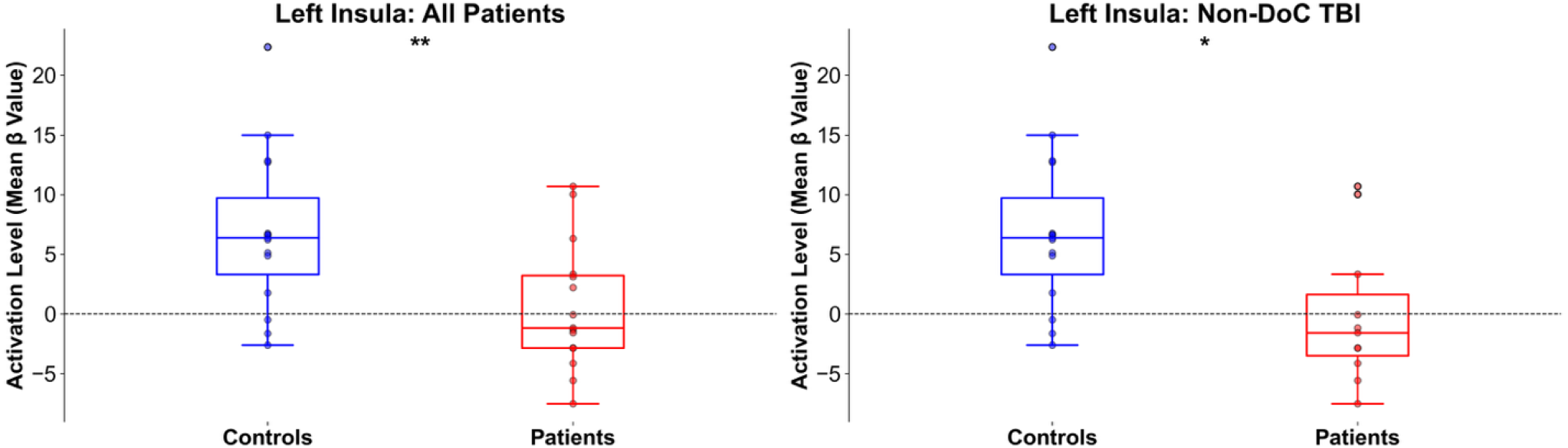
Box Plot of Activation Levels for Joy>Fear condition in the left insula. Points represent the difference in individual’s mean beta values for Joy>Fear conditions. Box limits represent the first and third quartiles, middle line represents the median, and whiskers represent the range of data within 1.5 times the interquartile range (IQR) from the lower and upper quartiles. * indicates a significance of p <0.05, ** indicates a significance threshold of p < 0.01.

### Resting-State Network Connectivity

Resting-state fMRI analyses were performed to explore whether there are disruptions in intrinsic connectivity within emotion-related networks. Seed-based whole-brain analysis showed significant differences between TBI and healthy controls in salience network connectivity with the caudate nucleus and the left hippocampus (p<0.001) (Fig 5A and B). These clusters remained significant when removing DoC TBI patients from the analysis. In contrast, no significant differences in connectivity of the DMN were found between TBI versus controls. Individual TBI patient Fisher transformed Z-values are plotted for each significant cluster (Fig 5C and D).

**FIGURE 5.**
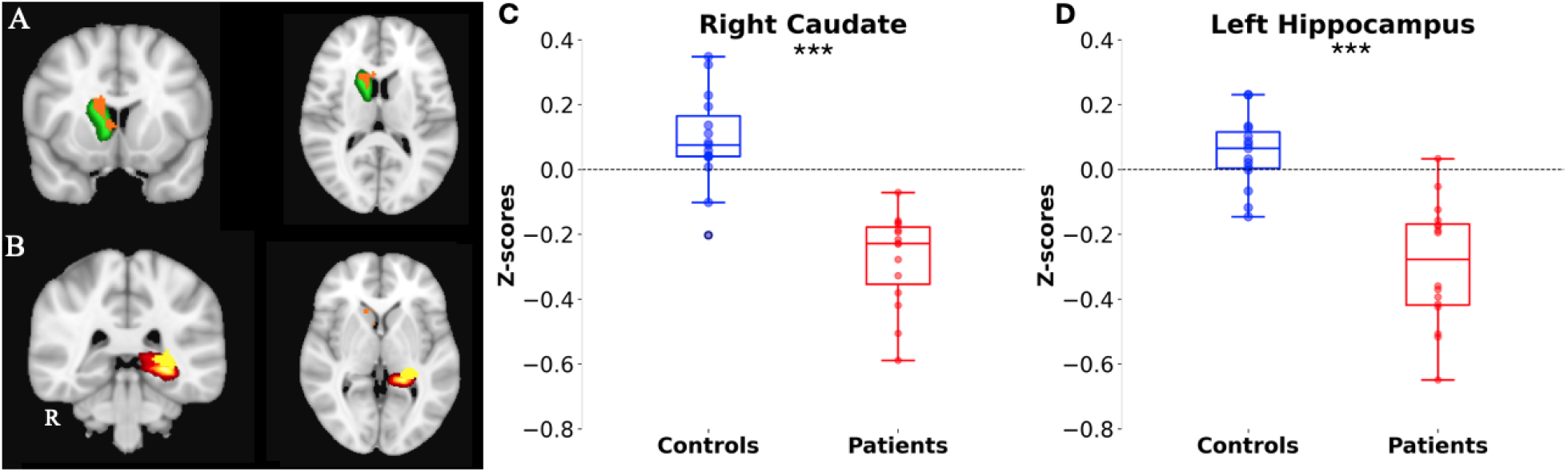
Salience network connectivity in seed-based analysis of healthy controls compared to TBI patients (n=16). (A) Orange cluster denotes higher connectivity between SN and the right caudate in healthy controls compared to TBI patients, centered at MNI coordinates (+14, +10, +18), overlayed on the right caudate nucleus from the Harvard Oxford cortical atlas (green). (B) Yellow cluster denotes higher connectivity between SN and the left hippocampus (centered at MNI coordinates −28, −32, +00) overlayed on the anatomical ROI (red). Images were overlayed on the MNI152 2mm standard template and were thresholder using clusters determined by Z>3.1 and a cluster significance threshold of p=0.05 (FWE corrected). (C) and (D) show the Fisher transformed Z values plotted as points for each group in the control > TBI contrast. Box limits represent the first and third quartiles, the middle line represents the median, and whiskers represent the range of data within 1.5 times the interquartile range (IQR) from the lower and upper quartiles. (C) Right caudate nucleus and (D) Left hippocampus. *** indicates a significance threshold of p<0.001.

#### Reduced mPFC–Amygdala Connectivity in TBI

Exploratory ROI-to-ROI connectivity analyses were conducted across all seed regions within and between the SN and DMN and key limbic regions (left and right amygdala). Significantly reduced connectivity was observed in TBI, compared to controls, for the following pairs of ROIs: mPFC and the right amygdala (T(28)=4.37, p<0.001), mPFC and the left amygdala (T(28)=3.09, p=0.005) (Fig 6).

**FIGURE 6.**
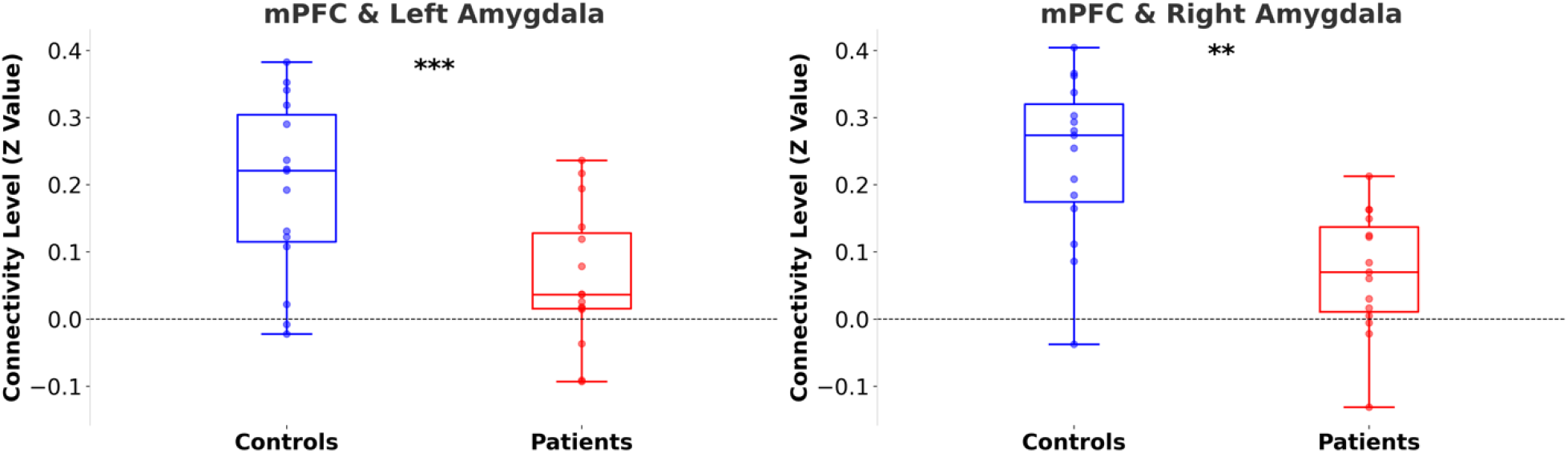
Resting State ROI-ROI Connectivity. Each plot shows the connectivity between two ROIs mPFC & Left Amygdala and mPFC & Right Amygdala. ** indicates a significance threshold of p < 0.01, and *** indicates a significance threshold of p < 0.001.

This finding remained significant after removing DoC TBI patients from the analysis. We found significantly lower connectivity for TBI patients between the mPFC and the right amygdala (T(27)=4.33, p<0.001) and between the mPFC and the left amygdala (T(27)=2.94, p=0.007), in keeping with the whole group results.

### Blunted Dark-Adaptation Response Suggests Autonomic Dysfunction in TBI

There was significantly greater pupillary dilatation, given by higher dark amplitude scores, in controls compared to TBI (F(1,24)=27.4, p<0.001), suggestive of sympathetic dysfunction in TBI. PLR latency was longer in TBI, compared to controls, at borderline significance (F(1,24)=3.92, p=0.060). In the light protocol, healthy controls and TBI patients did not show significant differences for PLR latency (F(1,24)=0.05, p=0.817), constriction velocity (F(1,24)=0.45, p=0.506), dilation velocity (F(1,24)=0.63, p=0.434), or amplitude (F(1,24)=0.489, p=0.491). We were unable to test the relationship between pupillometry and fMRI measures due to low numbers of complete datasets (n<10).

### Lower Depression Scores but Normative Anxiety Levels in Conscious TBI Patients

On the Neuro-QOL depression scale, conscious TBI patients had a mean T-score of 44.5 ± 6.04, which is statistically lower than the standardized population mean of 50 (t[12] = -3.30, p = 0.006). Because lower scores indicate fewer depressive symptoms, this reflects a slightly better mood than the population average; however, the difference is within 1 SD and is unlikely to be clinically meaningful. The mean anxiety T-score (47.44 ± 6.34) did not differ significantly from the standardized population mean of 50 (t[12] = -1.57, p = 0.142).

## Discussion

We examined the neural underpinnings of emotion processing in chronic survivors of severe TBI using stimulus-based fMRI, resting-state fMRI, and pupillometry. We found abnormality in activation of the left posterior insula during differential processing of joy compared to fear stimuli in individuals with chronic severe TBI compared to controls. Further, TBI was associated with altered resting-state SN connectivity with the caudate and hippocampus. These results were not attributable to differences in consciousness level. We also found diminished pupillary responses during dark-adaptation in TBI. Given the limited number of complete datasets, we were not able to test associations between autonomic dysfunction and emotion-related brain responses. Nevertheless, taken together, our findings indicate that diminished brain responses to emotional stimuli, disrupted brain connectivity, and impaired autonomic function are possible contributors to altered emotional processing in individuals with chronic severe TBI.

Our observation that posterior insula (pINS) responses are diminished in response to joyful relative to fearful stimuli in chronic severe TBI builds upon prior evidence for functional alterations in this region of the SN. The pINS has previously been found to differentially respond to music stimuli depending on the emotional valence.^15,33,39^ Attenuated activated of the pINS in TBI patients relative to healthy controls was not accompanied by similar findings in the right insula, or the ACC, suggesting lateralisation and neuroanatomic specificity in altered brain mechanisms underlying emotional processing in TBI. The pINS is termed the “primary interoceptive cortex”.^34^ Interoception, which is our sensing of internal body signals, is a vital process in normal emotional processing and experience.^40^ Therefore, our findings may reflect a specific deficit in interoception that contributes to the emotional processing of music after TBI. Findings from limited prior studies suggest that frontal lobe injuries (whether traumatic or not) are associated with reduced cardiac interoception,^41^ but a systematic exploration of interoception after severe TBI is needed.

Another possibility for the lateralized, region-specific findings may be the role of the insula in autonomic function. The left pINS is associated with sympathetic regulation, as identified through meta-analysis of fMRI studies investigating brain regions associated with high frequency heart rate variability and electrodermal activity.^42^ Of note, our pupillometry results suggest sympathetic dysfunction in the TBI cohort. Due to limited numbers of complete datasets, we are unable to test for a correlation between our pupillometry and fMRI data. Thus, the associations observed here do not provide causal evidence to mechanistically link autonomic dysfunction to altered emotional responses in chronic severe TBI. Nevertheless, our parallel findings of sympathetic dysfunction and abnormal neural processing of emotional stimuli in brain regions associated with sympathetic regulation in chronic severe TBI is motivation for further exploration to determine whether the observed blunting of dark-induced dilation may reflect dysfunction in central autonomic circuits^43^ that are relevant to emotional function. For example, it is possible that reduced sympathetic responsiveness may contribute to blunted affect and restricted affective reactivity reported some in individuals with chronic TBI.^4,19,44^ Only one small study has previously reported clinically relevant sympathetic and parasympathetic dysfunction in chronic TBI, and it did not explore relevance to emotional function.^19^ Future studies should seek to understand the shared brain mechanisms linking post-TBI autonomic and emotional dysfunction, as identification of commonly affected neuroanatomic regions may present an opportunity for novel neuromodulatory approaches.^45^

Our finding of reduced left pINS activation to joy versus fear stimuli in TBI remained significant after accounting for patients in DoC, suggesting that the finding is robust and not driven by altered arousal. Few studies have examined the capacity for emotional processing in this population.^46,47^ This highlights a gap in knowledge with substantial implications for understanding social interactions and optimizing quality of life for individuals with chronic DoC. Future studies are needed to identify patterns of disconnection within affective networks, such as the SN, that are still amendable to engagement by emotionally salient stimuli and that are physiologically receptive to therapeutic intervention.^48^

We also found reduced resting-state connectivity between the salience network and the right caudate and left hippocampus. While the caudate is not traditionally considered a limbic brain region, both the caudate and hippocampus are represented in minor clusters of an fMRI meta-analysis identifying brain regions important for processing of music-evoked emotion.^15^ The decreased SN-hippocampal connectivity may reflect the role of the hippocampus in integrating emotional salience with memory representations and attachment learning.^15,49^ The caudate is also implicated in dopaminergic signaling and cognitive control, and its involvement here may reflect disruptions in affective motivation or context-updating processes. Prior work has also demonstrated altered caudate connectivity in cognitive domains following TBI,^50^ suggesting that caudate functional connectivity may be multi-operational, contributing to both cognitive and affective processing. Moreover, the SN is important in both emotional processing and development of neurobehavioral problems.^51^ Additionally, TBI patients with depression have connectivity abnormalities of the dorsal attention network (DAN), a network with anatomical and functional overlap with the SN,^52^ suggesting that SN connectivity changes post-TBI may be clinically relevant. Caution is warranted in over-interpreting these resting-state findings in the absence of external stimuli, but our findings are consistent with prior observations that post-TBI network disconnection involves both cortical and subcortical networks.^50,53^

TBI patients in our cohort reported low depression scores and no detectable signs of anxiety. This does not necessarily reflect intact emotional function, as several severely impaired individuals were unable to complete the self-report. Additionally, alexithymia, which has been reported but poorly explored post-TBI,^54^ may contribute to impaired insight into one’s own emotional function. Further, incongruity between subjective ratings and objective impairment has been previously documented in TBI populations,^55^ which may reflect impaired insight or lower expectations around the extent of recovery after more severe injuries.

### Strengths & Limitations

The use of emotionally evocative music stimuli enhanced ecological validity, while the inclusion of severely injured patients extended our findings beyond the mild/moderate TBI literature. The inclusion of patients with a range of outcomes (including those still in DoC) increased the potential generalisability of our findings, however this choice comes with limitations. For example, we were not able to use visual stimuli (e.g., pictures or faces) to assess other aspects of emotional processing, nor were we able to explore active emotional processing, such as identification of emotional expressions. Furthermore, without active behavioral assessments of emotional function on the full sample, we cannot determine if or how our neuroimaging findings relate to real-world neurobehavioral problems post-TBI.

Owing to the convenience sampling approach to recruitment, which included patients at the low end of the recovery spectrum, a significant limitation of our study is the relatively small sample size and incomplete data availability across modalities. This meant we could not carry out our intended investigations into how the sympathetic dysfunction and brain activation and connectivity changes related to one another. Moreover, our cross-sectional design restricts causal inferences about recovery trajectories or longitudinal evolution of emotional dysregulation. Therefore, although we report one of the first studies to investigate mechanisms underlying emotional processing in chronic TBI, our results must be interpreted in the context of their preliminary nature.

## Conclusion

We found alterations in brain responses to emotional stimuli, SN connectivity, and autonomic function in individuals with chronic severe TBI. While these findings were not directly linked within individuals, they point to parallel disruptions that may contribute to the broader clinical picture of post-traumatic emotional and physiological dysregulation. Our findings also highlight the value of a multimodal approach to understanding post-TBI neurobehavioral problems, with implications for both research and clinical practice. Future studies should incorporate longitudinal designs, active emotional processing tasks, and larger sample sizes to better delineate mechanisms and inform targeted interventions.

## Acknowledgements

We thank the individuals who participated in the study, and their families. We are also grateful to the MRI technologists at Massachusetts General Hospital who contributed to data acquisition.

## Data availability

Data and code are available from the corresponding author upon reasonable request.

