## Supporting Information for "Emotional Processing After Severe Traumatic Brain Injury: Insights from Functional MRI and Pupillometry"

**Supporting Methods:**

**MRI Acquisition**

MRI data were acquired with a 32-channel head coil on a 3 Tesla Skyra MRI scanner (Siemens Medical Solutions; Erlangen, Germany) located in the Massachusetts General Hospital Neurosciences ICU. High-spatial resolution 3D T1-weighted multi-echo magnetization prepared gradient echo (MEMPRAGE) anatomical images were acquired for registration purposes. A blood-oxygen-level-dependent (BOLD) sequence was used for resting-state fMRI (12 minutes10 seconds) and for stimulus-based fMRI (5 minutes 54 seconds) with the following parameters: echo time (TE) = 30 ms, repetition time (TR) = 1250 ms, in-plane resolution = 2.0 x 2.0 mm, slice thickness = 2 mm, interslice gap = 2.5 mm, matrix = 954 x 954, field of view = 1908 x 1908 mm^2^, 72 slices, 4 factor simultaneous multi-slice acquisition. During resting fMRI, participants were instructed to keep their eyes closed. The first 19.5 seconds of data were used to obtain a stable baseline BOLD signal and an additional 2.5 seconds of data was used to optimize spatial co-registration, both excluded from further analysis. During stimulus-based fMRI, auditory stimuli were presented via MRI-compatible earphones (Newmatic Medical; Caledonia, MI) connected to the scanner’s sound system.

**Task design:**

Both runs followed identical durations, but the presentation order of music and rest blocks varied. Each run began with a 21.5s silence period to allow for stabilization of the BOLD signal. In each run, 4 fear and 4 joy stimuli were presented in a pseudo-random order, along with two rest periods (either 14.5s or 15.5s). No more than two stimuli from the same category were presented consecutively. The duration of each stimulus was 30.25s. The total run duration was 354s. Rest blocks were of varying lengths to ensure non-synchronization of stimuli blocks with the TR. All participants were presented with the same run order and were given varying breaks between scanning runs to cater to individual needs, typically a few minutes. To ensure consistency across both cohorts and to align with resting fMRI, participants were instructed to close their eyes. Music audios were presented both through loudspeakers in the scanner and MRI-compatible earphones (Newmatic Medical) at a volume level one notch above the level determined to be comfortable by fully conscious patients prior to MRI acquisition.

**Stimulus Based fMRI Preprocessing:**

The data was first converted from DICOM to nifti file format, then motion correction was applied using MCFLIRT to account for head movement during scanning.^1^ Skull stripping was then performed using the Brain Extraction Tool (BET) to remove non-brain tissue.^2^ Spatial smoothing was conducted with a Gaussian kernel of full width at half maximum (FWHM) of 5 mm to reduce noise and improve signal-to-noise ratio. Temporal filtering involved high-pass filtering with a Gaussian-weighted least-squares straight line fitting method (sigma value = 50.0s) to remove low-frequency artefacts. The data were then resampled and normalized to standard MNI152 space for consistency across subjects. Independent Component Analysis (ICA) was performed for each run using Multivariate Exploratory Linear Optimized Decomposition into Independent Components (MELODIC) Version 3.15.^3^ ICA-based automatic removal of motion artefacts (ICA-AROMA) was used to separate noise components, including motion-related components, from signal components,^4^ with manual checking of components to be removed by LML (experienced in fMRI analysis) prior to removal, following published guidance.^5^

Next, a first-level analysis was conducted to transform the data into a standard template space (MNI152) and convolve each fMRI run with a canonical hemodynamic response function Subject-level analysis was performed by constructing general linear models (GLM) with three regressors (anxiety, joy, neutral) convolved with a boxcar kernel model with canonical double-gamma hemodynamic response function (HRF) using FSL’s FEAT tool. Rest was not modelled in the GLM as a regressor, as it serves as an implicit baseline for the task-related conditions. Group-level analysis was performed to combine data from the two scanning runs for each participant and analyze mean activation across all participants on a whole brain level using FSL’s FMRIB's Local Analysis of Mixed Effects (FLAME 1+2) analysis.^6–8^

**RS-fMRI Preprocessing:**

Rs-fMRI data was analyzed using the CONN functional connectivity toolbox (release 22.a) in SPM (release 12.7771).^9,10^ Functional and anatomical data were preprocessed using a pipeline including realignment with correction of susceptibility distortion interactions, slice timing correction, outlier detection, direct segmentation, MNI-space normalization, and smoothing.^11^ Functional data were realigned using SPM realign & unwarp procedure^12^ and resampled using b-spline interpolation to correct for motion and magnetic susceptibility interactions. The artifact rejection tool was used to reject outlier scans that had a framewise displacement above 0.9 mm or global BOLD signal changes above 5 standard deviations.^13,14^ Functional and anatomical data were normalized into standard MNI space, segmented into grey matter, white matter, and CSF tissue classes, and resampled to 2 mm isotropic voxels.^15,16^ Functional data were smoothed using spatial convolution with a Gaussian kernel of 8 mm full width half maximum (FWHM). In addition, functional data were denoised using a standard denoising pipeline including the regression of potential confounding effects characterized by white matter timeseries (5 CompCor noise components), CSF timeseries (5 CompCor noise components), motion parameters and their first order derivatives (12 factors), outlier scans (below 285 factors), session effects and their first order derivatives (2 factors), and linear trends (2 factors) within each functional run, followed by bandpass frequency filtering of the BOLD timeseries between 0.008 Hz and 0.09 Hz.^11,13,17^ CompCornoise components within white matter and CSF were estimated by computing the average BOLD signal as well as the largest principal components orthogonal to the BOLD average, motion parameters, and outlier scans within each subject's eroded segmentation masks. ^18,19^

**Rs-fMRI seed based ROIs:**

Anatomical seeds were used to examine the default mode network (DMN) and salience network (SN), which were defined in CONN using Harvard-Oxford atlas.^20^ DMN: medial prefrontal cortex, right angular gyrus (LP), left LP, and posterior cingulate cortex. Salience Network: ACC, left anterior insula, right anterior insula, left rostal prefrontal cortex, right rostal prefrontal cortex, left supramarginal gyrus, right supramarginal gyrus. Following a commonly used method in prior studies of patients with disorders of consciousness (DoC),^21–23^ mean time series maps were generated by averaging the time series extracted from the four DMN (Fig 2B) seeds [1/4, ¼, ¼, ¼] and seven Salience Network (Fig 2C) seeds [1/7, 1/7, 1/7, 1/7, 1/7, 1/7, 1/7].

**Pupil examination**

To ensure the accuracy and reliability of the pupillometry measurements, participants underwent a pre-assessment protocol to rule out potential brain injury-related confounds that could have influenced pupil response. Participants were comfortably seated on a chair and asked to focus on a fixed point to maintain a consistent gaze throughout the measurement. The testing environment was controlled by positioning participants away from direct light sources or windows, with both corridor and room lights on to maintain consistent ambient lighting. Participants were instructed to avoid caffeine intake in the hours preceding the measurement to prevent potential confounding effects on pupil size.

Initially, ptosis was assessed by visually inspecting both eyelids to ensure they were symmetrically positioned above the pupils. Eye movements were then evaluated by asking the participant to follow a pen or finger moving in a cross (+) pattern approximately 30 cm from their face, which allowed testing of horizontal and vertical motion without head movement. To assess accommodation, participants were instructed to shift their focus from a distant point to a pen positioned close to the bridge of their nose, with expected pupil constriction indicating intact parasympathetic function. Direct and consensual light reflexes were tested by shining a light into one eye and observing for pupil constriction in the same (direct) and opposite (consensual) eye. This test was repeated for each eye, with careful positioning of the light source to avoid triggering an accommodative response. Relative afferent pupillary defect (RAPD) was then checked by swinging the light between eyes at a frequency of approximately 1 Hz to detect any abnormal dilation, potentially indicative of optic nerve damage. Finally, an ophthalmoscopy was performed to examine the optic disc in each eye, looking for differences in color and clarity; this was conducted with the lights dimmed to facilitate pupil dilation.

***Light and dark protocol testing procedures***

The light and dark stimulus protocols were used to assess the pupillary responses to changes in light conditions. The light protocol measured constriction in response to a brief light stimulus, while the dark protocol evaluated dilation during a period of darkness, providing insights into the function of both the autonomic and sympathetic nervous systems. For the light stimulus, single pulses of white light with a duration of 1 second at a bright intensity were delivered to induce a 30-40% constriction of the pupil. During each trial, the non-tested eye was directed to focus on a distant object (3-4 meters away) to maintain consistency across participants. The pupillometer was positioned over the tested eye, recording began with a 2-second baseline at 1mW, followed by a 1-second light pulse at 50mW, and then a 7-second recovery period at 1mW. This sequence was repeated three times for each eye, alternating between eyes with a 1-minute interval between trials. The dark stimulus protocol involved a step transition to darkness on a background of bright light. The non-tested eye was covered, and the participant was asked to focus on their reflection in the pupillometer lens to maintain gaze stability. The protocol began with a 2-second bright light at 121µW, followed by a 5-second dark step at 1µW, and concluded with another 2-second bright light at 121µW. Care was taken to ensure that the bright light did not produce any after-image effects, providing a clear assessment of the pupil’s response to changes in light intensity.

The light protocol involved presenting a brief light stimulus to the eye, and the PLR latency was calculated by measuring the time interval from stimulus onset to the beginning of pupil constriction. PLR constriction velocity was determined by calculating the rate of pupil constriction in mm/s during the constriction phase, while PLR dilation velocity was calculated by measuring the rate of pupil dilation in mm/s after the light stimulus was removed. Light amplitude was assessed by subtracting the minimum pupil size during constriction from the maximum pupil size before stimulus application. The dark protocol involved a period of darkness, during which PLR latency was measured by timing the onset of pupil dilation after light removal. Dark amplitude was calculated by subtracting the minimum pupil size during the dark adaptation from the maximum size observed following light removal.

**TBI Participant Details**

**TABLE S1.** Patient demographic and clinical characteristics

| Subject ID | Age | Sex | iGCS | Cause of TBI | Day of Scan Post Injury | LOC at Scan |
| --- | --- | --- | --- | --- | --- | --- |
| P1 | 18-29 | Male | 3 | MVA | 757 | FC |
| P2 | 60-69 | Female | 3 | MVA | 675 | FC |
| P3 | 18-29 | Male | 3 | MVA | 717 | FC |
| P4 | 18-29 | Male | 4 | MVA | 798 | FC |
| P5 | 18-29 | Female | 4 | Fall | 581 | FC |
| P6 | 18-29 | Male | 3 | MVA | 1892 | MCS |
| P7 | 30-39 | Female | 3 | Sport | 389 | FC |
| P8 | 60-69 | Female | 3 | MVA | 711 | FC |
| P9 | 18-29 | Male | 7 | Fall | 639 | FC |
| P10 | 70-73 | Male | 3 | MVA | 253 | FC |
| P11 | 18-29 | Female | 7 | MVA | 644 | FC |
| P12 | 30-39 | Female | 3 | MVA | 412 | FC |
| P13 | 18-29 | Female | 3 | Fall | 1853 | FC |
| P14 | 30-39 | Female | 3 | MVA | 560 | MCS |
| P15 | 18-29 | Female | 4 | MVA | 457 | VS |
| P16 | 30-39 | Male | 4 | MVA | 428 | FC |
| P17 | 30-39 | Female | 4 | MVA | 564 | FC |
| P18 | 60-69 | Male | 4 | MVA | 252 | PTCS |
| P19 | 18-29 | Male | 4 | MVA | 728 | MCS |
| P20 | 18-29 | Female | 4 | MVA | 2562 | MCS |
| P21 | 40-49 | Female | 4 | MVA | 370 | FC |
| P22 | 18-29 | Male | 4 | MVA | 375 | FC |
| P23 | 18-29 | Male | 4 | MVA | 473 | FC |
| P24 | 30-39 | Female | 4 | MVA | 194 | VS |
| P25 | 18-29 | Female | 4 | MVA | 3770 | VS |
| P26 | 30-39 | Female | 4 | Sport | 410 | FC |

GCS (iGCS) is defined as the initial post-resuscitation GCS score assessed by a qualified clinician who performed a reliable examination (not confounded by sedation and/or paralytics) prior to ICU admission. LoC is assessed at time of scan via behavioural evaluation with the CRS-R as coma, VS, MCS-, MCS+, PTCS (emerged from MCS but disoriented), FC = fully conscious. Abbreviations: MVA = motor vehicle accident.

**TABLE S2. Table depicting participant inclusion and exclusion across data types.** Table illustrates the number of participants for whom each data type was collected. All exclusions were for data quality reasons.

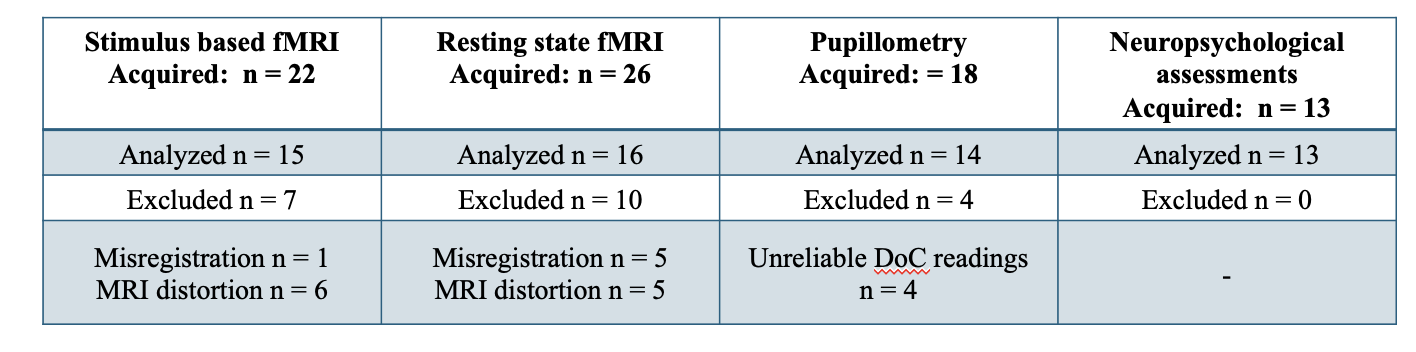

**TABLE S3.** Summary statistics of results from stimulus-based fMRI ROI analysis

| **Comparison** | **ROI** | **HC Mean ± SD** | **TBI Mean ± SD** | **Summary Statistics** |
| --- | --- | --- | --- | --- |
| Healthy control vs all TBI | Left Insula | 6.84 ± 6.69 | 0.58 ± 5.36 | T(28) = 2.83, p = 0.009 |
| Healthy control vs all TBI | Right Insula | 6.75 ± 6.84 | 3.69 ± 6.02 | T(28) = 1.30, p = 0.205 |
| Healthy control vs all TBI | Left Amygdala | 0.14 ± 3.29 | -1.23 ± 4.25 | T(28) = 0.98, p = 0.334 |
| Healthy control vs all TBI | Right Amygdala | -0.98 ± 4.45 | 0.57 ± 3.98 | T(28) = -1.01, p = 0.323 |
| Healthy control vs non-DoC TBI | Left Insula | 6.84 ± 6.69 | 0.28 ± 5.98 | T(24) = 2.58, p = 0.016 |
| Healthy control vs non-DoC TBI | Right Insula | 6.75 ± 6.84 | 3.28 ± 6.02 | T(24) = 1.34, p = 0.192 |
| Healthy control vs non-DoC TBI | Left Amygdala | 0.14 ± 3.29 | -0.98 ± 4.47 | T(24) = 1.06, p = 0.299 |
| Healthy control vs non-DoC TBI | Right Amygdala | -0.98 ± 4.45 | 0.86 ± 4.09 | T(24) = -1.08, p = 0.293 |

Mean values are calculated based on the beta values extracted from the contrast of joy>fear conditions.
